# Divergent avian strains drive an off-season influenza A peak in municipal wastewater

**DOI:** 10.64898/2026.04.02.26350079

**Authors:** Alexander L. Jaffe, Alessandro Zulli, Dorothea Duong, Bridgette Shelden, Miriam Goldman, Miles Richardson, Marlene K. Wolfe, Alexandria B. Boehm

**Affiliations:** Department of Civil and Environmental Engineering, Stanford University; Verily Life Sciences, South San Francisco, CA; Gangarosa Department of Environmental Health, Rollins School of Public Health, Emory University

## Abstract

Wastewater sequencing is an increasingly valuable tool in tracking the spread of infectious disease agents across space and time in areas of dense human settlement. Among pathogens that can be readily detected by this approach is influenza A, which follows predictable patterns of prevalence through the winter months in North America. Here, we leverage routine surveillance of a municipal wastewater treatment plant in Northern California to describe an atypical, off-season spike in influenza A concentrations that rivals that of the winter respiratory virus season. Drawing upon metagenomic data generated through hybrid-capture sequencing, we assemble and subsequently characterize fragments of divergent influenza genomes that appear to derive predominantly from the avian H16 clade. These strains exhibit close evolutionary relationships to influenza isolated from migratory shorebirds, hinting at potential host species and mechanisms of geographic spread. Analysis of read abundances suggest that these avian strains dominate the pool of influenza circulating during the summer months, when typical human-infecting strains are essentially absent. Together, our results expand the value of wastewater sequencing to encompass sensitive tracking of outbreaks within animals in interface regions where human settlement abuts wildlands, increasing overall pandemic preparedness.

## Introduction

Influenza A virus (IAV) is one of the leading causes of morbidity and mortality worldwide, and one of the major respiratory illnesses impacting human health.^1,2^ The World Health Organization currently estimates that IAV is responsible for up to 600,000 deaths and 1 billion cases each year.^1^ Influenza A is seasonal, with most cases occurring in each hemisphere’s winter season, and there is a vaccine available, though both immune history and antigenic mismatch can lower vaccine effectiveness.^3,4^ The majority of IAV cases each year are caused by commonly circulated IAV subtypes such as H3N2 and H1N1. However, IAV can undergo genetic reassortment and has broad host tropism^5^, allowing new subtypes and variants to emerge, sometimes with pandemic potential.

The most recent IAV pandemic was in 2009, with a quadruple reassorted swine influenza H1N1 infecting an estimated 11-21% of the global population and causing an estimated 575,400 deaths.^6^ This recent flu pandemic highlights how new variants emerge through zoonoses and spillover and emphasizes the need for surveillance of influenza evolution.^6,7^ Studying and predicting the evolution of influenza as it undergoes genetic reassortment through multiple hosts is key to preventing future pandemics, allowing researchers to rapidly develop vaccines and other countermeasures.^8,9^ Significant advances have been made in predicting IAV evolution, including using deep-learning and transformer based-models to predict viral fitness based on predicted sequences and protein conformation.^9,10^ These efforts require massive amounts of data, and to that end, a multitude of surveillance systems have emerged both nationally and internationally for sequencing and disseminating genome sequences, ranging from clinical (Center for Disease Control, Global Initiative on Sharing All Influenza Data) to environmental (Animal and Plant Health Inspection Service).^11–13^ These are all capital and time intensive undertakings that could be complemented by wastewater-based surveillance.

Recently, there has been significant progress in using wastewater surveillance as an integral tool for studying IAV epidemiology.^14–16^ These have focused on quantitative approaches like quantitative polymerase chain reactions (qPCR) or digital droplet polymerase chain reactions (ddPCR) that are limited to identifying either all types of IAV simultaneously, or specific types individually. Despite this limitation, this approach has been successfully used to track both human IAV and emerging avian strains such as H5N1.^17^ Since 2024, wastewater has been used in the United States to track H5N1 due to outbreaks in cattle and the resulting risk of spillover into human populations.^18,19^ Recently, a newer approach to wastewater-based surveillance of IAV, probe-based hybridization and sequencing, has demonstrated promising results for quantitative IAV surveillance while also allowing for simultaneous strain subtyping.^20^

In this study, we present the results of an investigation into an atypical peak in IAV wastewater concentrations comparable to the winter season in Palo Alto, California during the summer of 2025. We combine reverse-transcription ddPCR (RT-ddPCR) results with probe-based hybridization sequencing to systematically exclude common circulating IAV subtypes from humans, and identify a set of avian strains likely associated with migratory shorebirds as the main source of this viral material. This study demonstrates the value of metagenomic sequencing of wastewater samples to complement existing PCR-based surveillance systems, even for pathogens of non-human animals.

## Methods

### Sample collection and storage

One hundred and forty-seven wastewater treatment plants (WWTPs) located in 40 US states provided either “grab” samples from primary clarifiers or 24-hour composite samples of influent from the headworks (Table S1). Depending on WWTP location, samples were collected between 3-7 times a week between September 1, 2024 and November 1, 2025. Additional details of sampling as well as the location of each WWTP are available in Boehm et al.^21,22^ After collection, samples were then stored at 4°C, shipped on ice to the laboratory, and processed immediately for RT-ddPCR analysis. Aliquots of samples from Palo Alto WWTP were taken and stored at 4°C for ∼0.5-10 weeks (depending on sample) for later metagenomic sequencing. Storage temperature was informed by our previous work indicating that wastewater storage at 4°C is preferable over freezing, as freeze-thaw cycles can increase the degradation of nucleic acids.^23^

### RT-ddPCR quantification

Nucleic acids were extracted from wastewater solids and used neat as template in RT-ddPCR to quantify IAV genomes using commercial kits, as previously described.^17,21,22^ Briefly, wastewater solids were centrifuged at 24,000×g and resuspended at 75 mg/ml in a buffer containing bovine coronavirus (BCoV) nucleic acid recovery control. After homogenization and nucleic acid extraction from 6-10 replicate aliquots, RT-ddPCR was performed using established assays targeting the conserved M gene of IAV, as well as hemagglutinin (HA) gene subtypes H1, H3, and H5.^21^ IAV H3 concentrations were measured after March 2025 only.^21^ The 6-10 nucleic acid extract replicates were run in their own wells. Results required ≥3 positive droplets across merged wells for detection, corresponding to ∼500-1000 copies/gram. Full nucleic-acid extraction and quantification methods are provided in previous data descriptors.^21,22^ Methods followed EMMI and MIQE guidelines and thus included negative and positive controls, and replication as explained in Boehm et al.^21,24,25^

Concentrations of M, H1, H3, and H5 markers were reported as population weighted concentrations as described in Chan et al.^26,27^ Five-day trimmed averages were also calculated and plotted as described elsewhere.^26,27^

### Sample preparation for metagenomic sequencing

Biobanked Palo Alto wastewater samples were warmed at room temperature for 30-45 minutes before processing. Samples were spun down at 4200xg for 5 minutes, and 50 mL of resulting supernatant was vacuum filtered through 0.45 μm pore size polyethersulfone (PES) filters (Thermo Scientific #165-0045, Waltham, Massachusetts, USA). Viral particles were then concentrated from 10 mL of the filtrate using Ceres Nanotrap Microbiome A particles (Manassas, Virginia, USA). Total nucleic acids were then extracted using the MagMAX Viral/Pathogen Kit (Waltham, Massachusetts, USA) on an automated KingFisher instrument, followed by PCR inhibitor removal using the Zymo OneStep Kit. Nucleic acids were then diluted 1:10 with nuclease-free water for all samples except wwd006407 and wwd006408 (January 2026), which were prepared without dilution. cDNA synthesis was performed using a ProtoScript II First Strand cDNA Synthesis Kit (New England Biolabs #E6560L, Ipswich, Massachusetts, USA). cDNA was subsequently purified using Kapa HyperPure Beads, and samples were normalized to a concentration of 1 ng/μL using nuclease-free water for library preparation. Enzymatic fragmentation, adapter ligation, and PCR amplification were accomplished using the Twist Library Preparation Kit (Twist Biosciences, #103548). Libraries were additionally prepared with the Twist Unique Molecular Identifiers (UMI) adapter system.

Finally, libraries were enriched using the Twist Comprehensive Viral Research Panel (Twist Biosciences, #103548), which targets influenza A genomes among more than 3000 viral genomes, through hybridization at 70°C for 16 hours. Ribosomal RNA was depleted using CRISPR-Cas9 technology (Jumpcode Genomics, #KIT1014). Prepared libraries were ultimately sequenced on an Illumina NovaSeq X Plus (150 or 250 bp paired-end reads), alongside negative extraction/sequencing controls and a positive control, consisting of ATCC virome nucleic acid mix (MSA-1008) at a concentration of 1000 genome copies per reaction mixed with 10 ng *E. coli* genomic DNA (Zymo D5016) and 10 ng *E. coli* total RNA (Invitrogen AM7940). Full library preparation details are provided on protocols.io.^28^

### Sequence read processing and assembly

Sequencing reads were screened for those of human origin using a three-step, iterative process employing Kraken2 (v2.1.3), bowtie2 (v2.5.2) with reference GRCh38.p14, HRRT (v2.2.1) with the human_filter.db.20250916v2 database. Reads were next trimmed of sequencing adapters and UMIs using fastp (v0.23.4), and de-duplicated using HUMID (https://github.com/jfjlaros/HUMID).^29^ Filtered and de-duplicated reads were subjected to initial classification with EsViritu v1.1.0.^30,31^ Read profiling was complemented by *de novo* assembly of full hybrid capture metagenomes. Specifically, each set of sequenced reads were subjected to assembly by SPAdes in RNA viral mode (default parameters).^32,33^ To identify assembled influenza segments, assembled contigs were BLASTed against all influenza segments contained within the EsViritu database (database v3.2.2). Any contig that attained ≥50% coverage when aligned against a reference segment was retained.

Putative influenza segments were combined with reference influenza genomes from the EsViritu database as well as H16 genomes drawn from the NCBI (-nt database) and GISAID (https://gisaid.org/), including genomes from several recent studies.^34–37^ A full table of the metadata of sequences from GISAID is available in the Supporting Information (Table S2). Each segment was stored as a separate FASTA record, and the total sequence set was clustered using USEARCH at 95% and 99% identity (https://www.drive5.com/usearch/) (*-cluster_fast)*. Clusters were filtered to those corresponding to the HA and neuraminidase (NA) segments (segments 4 and 6, respectively), and the longest contig within each cluster was selected as a representative. Where clusters contained a newly-assembled wastewater fragment, the longest wastewater fragment was chosen as a cluster representative. To ensure that representative fragments did not contain assembly errors, de-duplicated sequencing reads were aligned to representative fragments using bowtie2 and assessed manually for regions without read support. One such alignment is visualized in Fig. S1.

### Phylogenetic reconstruction of assembled sequences

To reconstruct the identity and evolutionary history of newly-assembled flu fragments, both protein and nucleotide trees were constructed for the HA and NA segments, respectively. For protein trees, cluster representatives at 95% nucleotide identity were concatenated, and encoded protein sequences were predicted using Prodigal (*-p meta*).^38^ Proteins were aligned using MAFFT (*--auto*) if they exceeded 100 amino acids in length, achieved ≥50% of the total alignment length, and contained fewer than 10 ambiguous bases.^39^ This filtering process removed one HA sequence from consideration (WWD005552_RVS_scaffold_2984), possibly due to ambiguities in sequence assembly or errors in the prediction of open reading frames. Alignments were subsequently trimmed using trimal (*-gt 0*.*1*) to remove uninformative positions.^40^ Finally, the protein tree was inferred using IQTree2 with a model selection step (*-bnni -m TEST -st AA -bb 1000 -nt AUTO)*, which selected FLU+I+G4 as the best-fitted model.^41^

For the HA segment, a more detailed nucleotide tree was then constructed using all sequence clusters in the immediate phylogenetic context of wastewater contigs, as well as a nearby outgroup. For this tree, formerly redundant sequences were restored for greater phylogenetic resolution, as long as these sequences obtained at least 300 nucleotides in length. Otherwise, the same bioinformatic steps were employed. Host type, geographic location, and year of collection were derived from each database, manually curated where missing, and visualized alongside the tree using iTol.^42^

We also undertook additional analyses of sequence similarity for the HA segment. All redundant sequences from clusters that were phylogenetically proximal to wastewater contigs were included, provided that these contigs attained 1000 bp in length or greater. Comparisons were then made between these sequences and wastewater contigs by first predicting nucleotide gene sequences with prodigal, aligning and alignment trimming (as above), and calculation of pairwise percent identity across the trimmed alignment at all non-gapped sites. Preliminary subtype classifications for segments other than HA/NA were made by subjecting reconstructed nucleic acid to BLASTn (core_nt database) and selecting the top hit by total bitscore.

### Temporal dynamics of influenza subtypes

To study abundance dynamics of influenza subtypes over time, we examined the recruitment of metagenomic reads by representative HA and NA sequences from both our *de novo* assemblies as well as public data repositories. Cluster representatives (95% identity) were concatenated and used to construct a bowtie2 index. Next, trimmed and de-duplicated metagenomic reads were competitively aligned to this index, also using bowtie2.^43^ For each alignment file, reads aligned to representative sequences were identified using the fetch function in pysam (https://github.com/pysam-developers/pysam). Individual reads (forward or reverse) were classified as ‘stringently mapped’ if the number of mismatches between read and reference was less than or equal to 5% of the read length after trimming. Otherwise, reads were classified as ‘loosely mapped’. The proportion of reads mapping both stringently and loosely to each representative sequence was then visualized across the time series as a function of either HA or NA clade. Finally, read counts for each segment were summed as a function of collection date and normalized by the total number of trimmed reads in the sample to calculate effective sequencing depth in reads per million (RPM). All numerical data were plotted and analyzed using RStudio (v4.5.1) and the seaborn library in Python.

## Results and Discussion

### An off-season peak of Influenza A in Palo Alto municipal wastewater

RT-ddPCR results showed a large influenza A spike in measurements of the M gene beginning in July of 2025. This spike was unique to Palo Alto and not present nationwide (146 other WWTPs). **Figure 1** shows how atypical these measurements were, with very few detections nationwide during that period, while Palo Alto showed sustained high detections ≥3,000 gene copies per gram dry weight (Table S3, S4). Investigating these detections using HA subtype specific RT-ddPCR assays to detect circulating H1, H3 and H5 sequences yielded only sporadic detections which did not adequately explain the high levels of IAV M gene.^22^ Notably, H1, H3, and H5 were also not detected at substantial concentrations at other WWTPs in the study (Figure 1). All assays were confirmed to be sensitive and specific to contemporary strains based on *in silico* analyses (Supplementary Methods).

**Figure 1.**
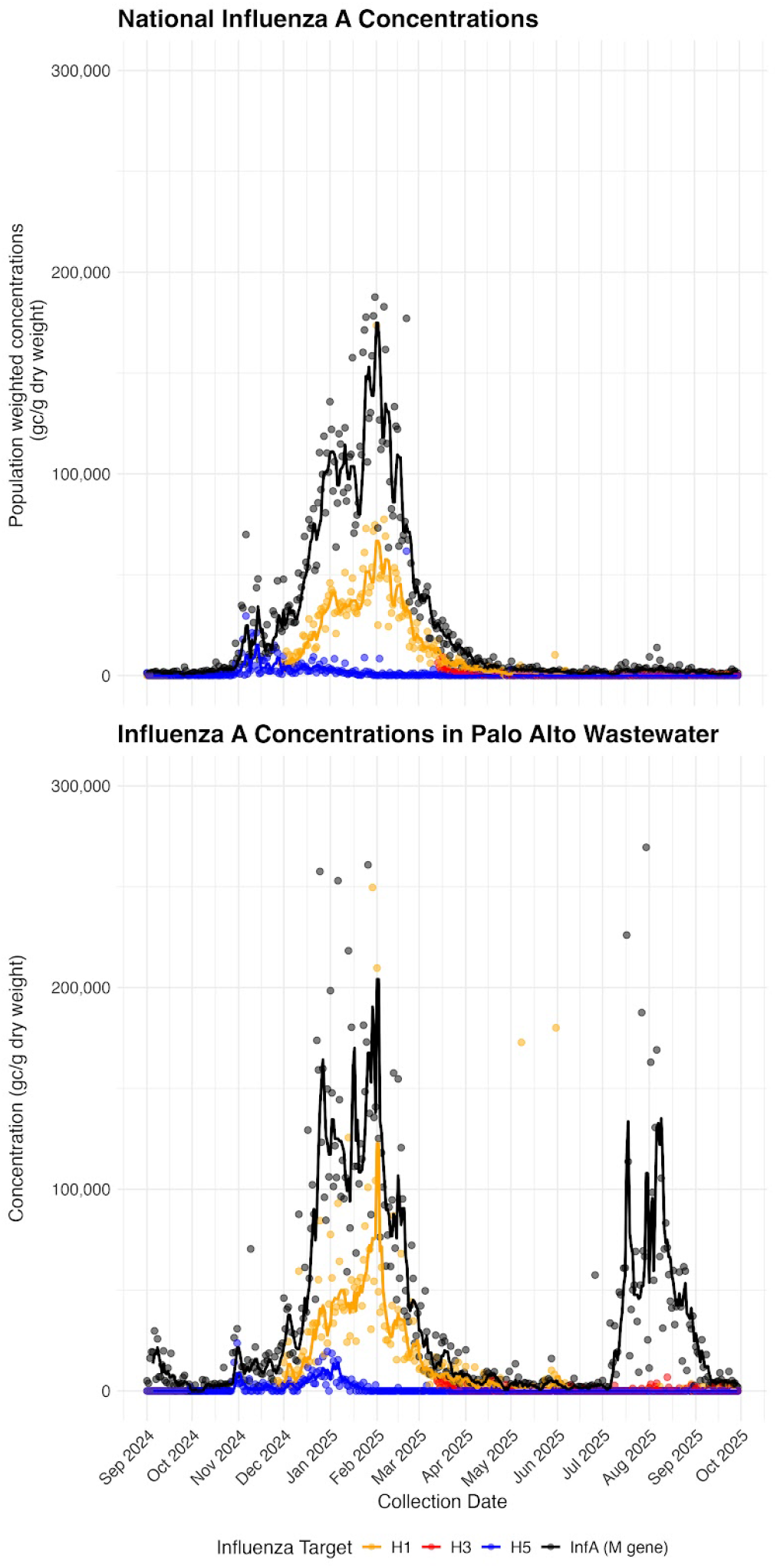
(Upper panel) Population weighted average national concentrations of influenza A M, H1, H3, and H5 genes in wastewater across 147 WWTPs, including Palo Alto, in 40 states between September 2024 and November 2025. Dots represent population weighted concentration in gene copies per gram of dry weight while lines represent 5-day trimmed averages. (Lower panel) Concentrations of influenza A M, H1, H3, and H5 genes in wastewater in Palo Alto, California. Dots represent concentration in gene copies per gram of dry weight while lines represent 5-day trimmed averages.

### Influenza A sequences assembled from wastewater likely derive from shorebirds

To more deeply investigate the biological basis of this pattern, we performed hybrid capture sequencing targeting a wide variety of animal viruses (including influenza A) on 14 daily samples of wastewater solids from the Palo Alto WWTP, corresponding to the highest IAV concentrations observed during the summer peak (Methods). Resulting sequencing reads were initially analyzed using a read-based profiler (Methods), which indicated that a mixture of influenza subtypes - including H13N6, H7N7, H4N8, and H6N5 - was detected based on read mapping to reference genomes (Table S5). However, recruited reads only attained ∼94.9% identity to reference sequences on average, suggesting the potential presence of divergent strain(s) (Table S5). Read profiling of negative extraction and sequencing controls yielded only detections of human adenovirus and papillomavirus (Table S5).

To evaluate the potential presence of divergent influenza strains, we performed *de novo* assembly of each set of metagenomic reads, utilizing an algorithm tailored for RNA viruses (Methods). Despite low sequencing depth, we successfully assembled contigs that appeared to derive from all eight segments of the influenza genome, including the ‘typing’ segments encoding the hemagglutinin and neuraminidase proteins (Table S6). Clustering of assembled contigs at 95% identity indicated that between 3 and 5 distinct clusters were assembled per segment, suggesting that some degree of sequence diversity was present among influenza strains (Table S6). We next focused on the HA (max length 1,743 bp) and NA segments (max length 1,478 bp), predicting their encoded proteins and placing them within a phylogenetic tree alongside sequences from diverse reference strains (Fig. 2a, Fig. S2). Intriguingly, assembled HA proteins fell within a clade derived from H16 (primarily H16N3) – an avian subtype primarily hosted by gulls and terns – in contrast with our initial read-based analysis.^36,44,45^ A more detailed tree based on HA gene nucleotide sequence confirmed this placement and further revealed that assembled sequences were closely related to one another, forming a monophyletic group within a larger clade of strains derived primarily from North American gulls (Fig. 2a).

**Figure 2.**
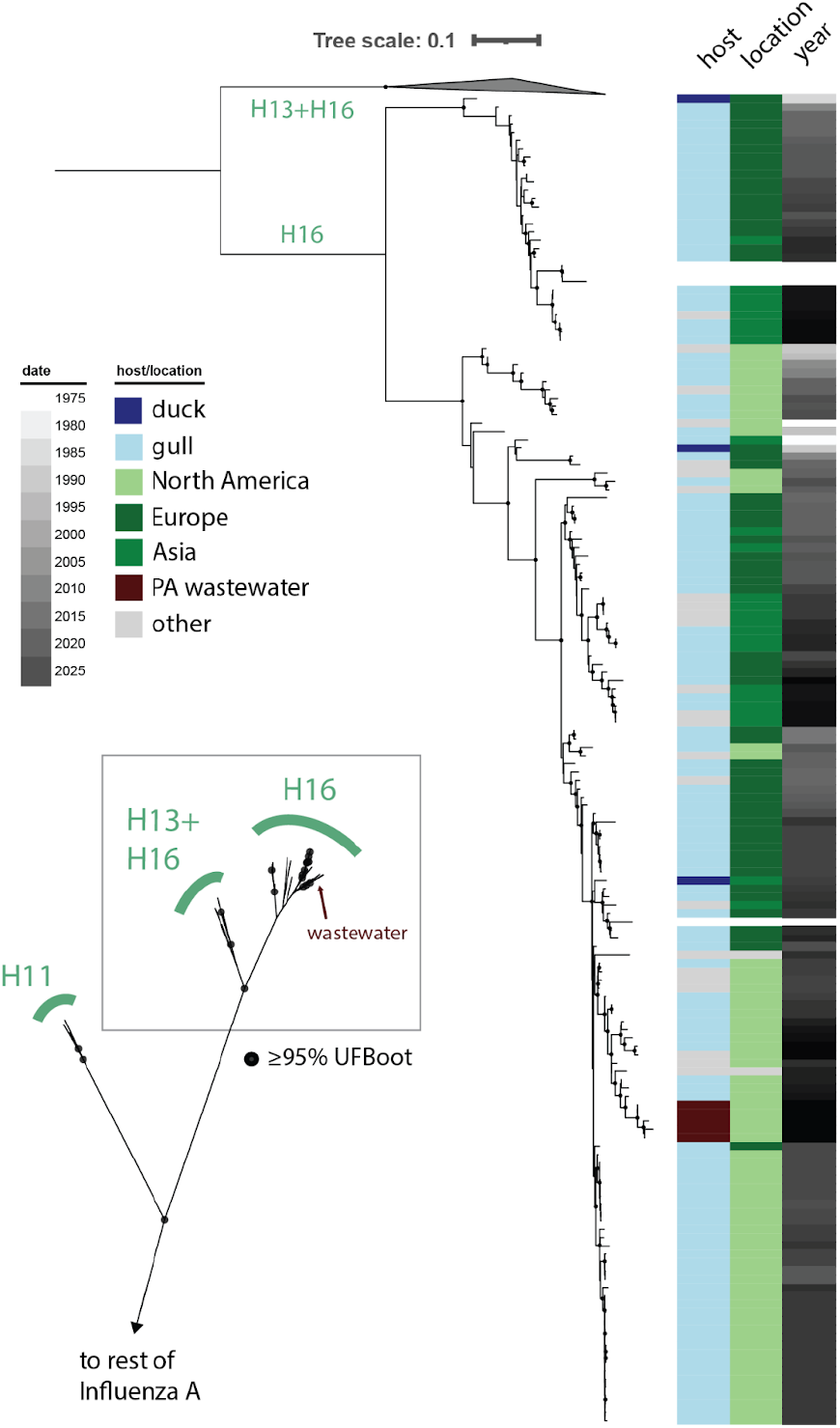
Phylogenetic trees depicting evolutionary relationships of wastewater strains to reference sequences based on HA protein sequence (inset) and HA nucleotide sequence (main tree). In both trees, black dots represent nodes supported by ≥95% ultra-fast (UF) bootstrap.

The nucleotide tree topology also revealed that an H16N3 strain sampled from an Alaskan gull in 2021 (EPI_ISL_16641800, A/gull/Alaska/21MB00111/2021) was the closest relative of our newly-reconstructed HA genes. Analysis based solely on sequence similarity, rather than evolutionary reconstruction, suggested that this gene obtained ∼96.1% identity to one of our newly-assembled genes, the most similar of any comparison made with reference sequences (Table S7). Other newly-assembled HA genes were substantially more divergent from reference sequences, attaining only 74.4%-82.9% identity at most (Table S7). To our knowledge, no more closely related strains have been sequenced to date, possibly due to lack of sampling effort on the U.S. western coast after 2021. We anticipate that sequencing of historical samples from these bird populations would reveal currently missing links between the wastewater strains presented here and their closest relatives in the public record, which emerged 4-5 years prior.

Phylogenetic placement of assembled NA proteins, on the other hand, revealed two sequences falling in with N3 (again, mostly H16N3) references (Fig. S2). We also recovered one sequence that fell within the N8 clade, which was most closely related to diverse avian and swine N8 strains. With the methods employed here, it remains unclear whether this recovered N8 fragment was associated with an HA segment from the H16 clade, or another that could not be assembled. However, given the repeated assembly of HA segments from the H16 clade, as well as NA segments from the N3 clade, we consider the presence of at least one H16N3 strain in sampled wastewater to be highly likely. This inference is consistent with patterns of sequence similarity observed for other segments assembled but not subjected to phylogenetic reconstruction, which mostly resembled H16N3 strains (Table S6).

### Abundance dynamics and potential host range of wastewater influenza strains

Finally, to assess the extent to which the identified strains could drive the unusual influenza peak observed in the RT-ddPCR data (Fig. 1), we examined abundance patterns of strains across the time series. By competitively recruiting reads to the set of HA contigs drawn from our assemblies as well as existing references from diverse influenza subtypes, we observed that H16 sequences were dominant through the late July/August peak (Fig. 3, Table S8). Newly-assembled H16 segments recruited reads at all time points where HA was detected. Furthermore, most reads were mapped stringently (Methods), suggesting that mapping targets were generally representative of the sequence diversity present. In one instance, where effective sequencing depth was greater, we also detected low relative abundance of two avian H3 strains (Table S8). Supporting the robustness of our method, application of the same approach to additional samples from January 2026 revealed a read population entirely attributable to mammalian H3 strains (Fig. S3, Table S8). These findings are consistent with clinical reports showing that H3N2 was the dominant IAV subtype circulating among humans in the winter 2025-2026 season.^46^ In several instances, no HA reads were detected in wastewater samples, likely due to variable efficiency of hybrid capture (‘n.d.’).

**Figure 3.**
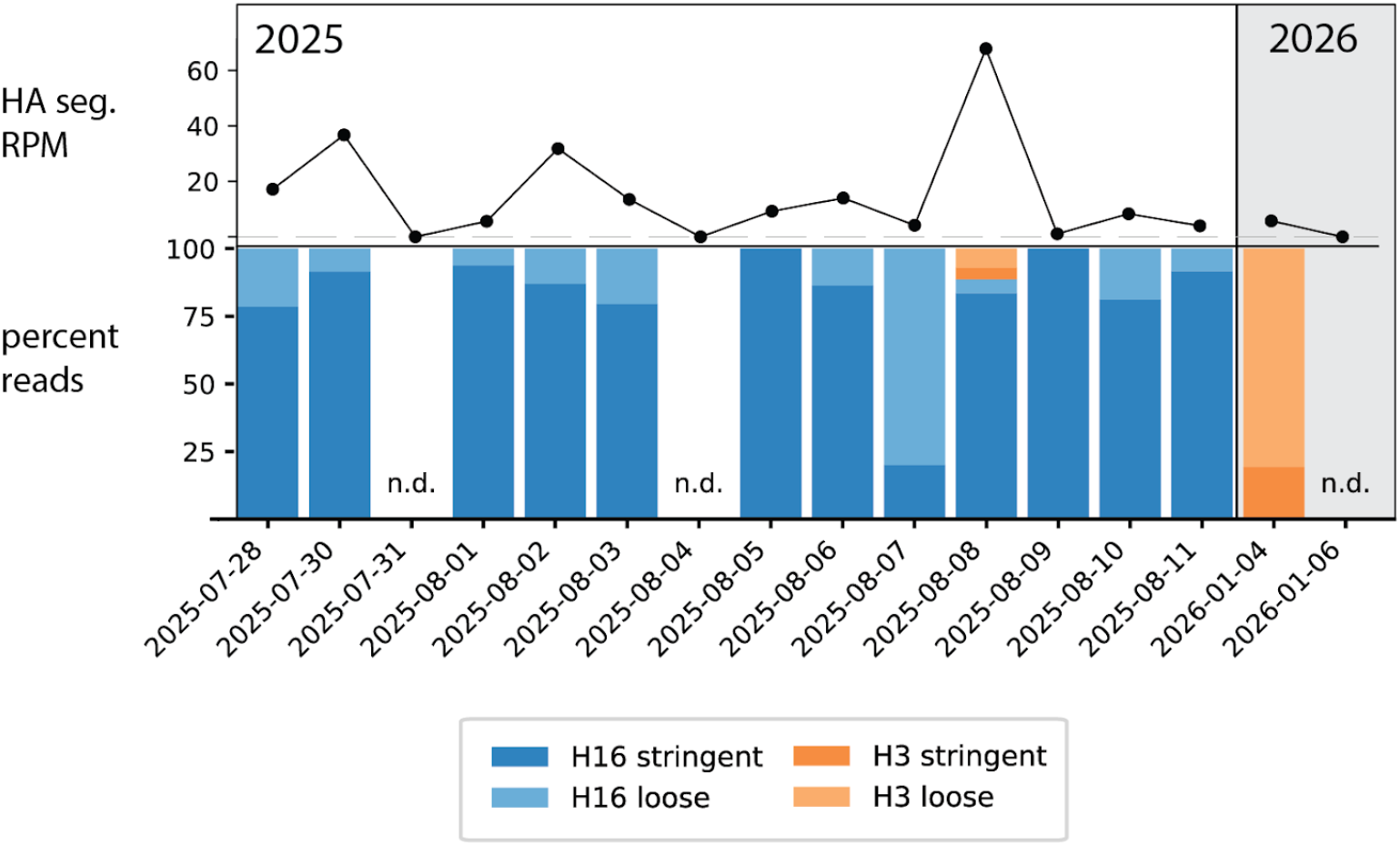
Relative abundance dynamics of wastewater strains across the time series, based on reads mapped to a database of HA segments, including newly-assembled sequences. Reads mapping stringently to HA segments, as well as those mapping loosely (Methods) are depicted in different shades. Upper panel indicates the total number of HA reads considered at each time point, expressed in reads per million (RPM). Abbreviations: HA, hemagglutinin; n.d., not detected.

We applied this same approach to study strain dynamics based on the NA segment (Figure S2). There, we observed that N3-type genes accounted for most reads over time, whereas N8 genes only appeared to be numerically relevant at one time point (Fig. S2, Table S8). Overall, we infer that H16 and N3 type strains are primarily responsible for the observed wastewater peak, though we cannot definitively rule out the presence of other, low-abundance strains that were unable to be assembled here or that are otherwise absent from reference databases.

Regardless of the precise subtypes in circulation, our combined results raise intriguing questions about the sources of viral material sampled from wastewater. Palo Alto WWTP is situated less than 2 kilometers away from a large nature preserve (Fig. S3) comprising nearly 2,000 acres of pristine marshland and hosting large numbers of both resident and migratory shorebirds.^47^ While input of environmental runoff is theoretically rare in ‘closed’ WWTP systems like this one, we speculate that standing water from the preserve may enter into the system through a currently unknown mechanism; or, alternatively, fecal material may directly enter the system via recirculation of effluent from open-air settling tanks, where shorebirds can congregate.^48^ One factor to consider in evaluating these hypotheses is that many infected individuals would likely be required to achieve the influenza concentrations observed by RT-ddPCR, although these estimates are sensitive to shedding rates used for modeling.^48^

Of course, the composite nature of wastewater does not permit definitive identification of host species for the strains analyzed here. However, our molecular evidence points firmly to a closely related set of H16Nx strains from a larger clade hosted primarily by gulls (Fig. 2). Furthermore, the timing of the influenza peak observed in Palo Alto wastewater is highly consistent with previous observations that IAV prevalence is highest among gulls during the late summer and early fall, potentially coinciding with the end of the breeding season for some Northern hemisphere species.^45^ During this time, both adults and immunologically naive juveniles aggregate into dense colonies that may greatly favor the spread of viruses.^45^ Thus, the wastewater peak observed in our study could reflect the dynamics of an IAV outbreak among avian hosts, rather than a temporary incursion of fecal material into wastewater streams. In the future, sampling of birds during similar anomalies will both allow for more clear delineation of host range as well as the reconstruction of genome sequences for individual strains, complementing the population-level consensuses generated here with metagenomics.

Finally, the finding that wastewater IAV sequences were most closely related to a strain detected nearly 4-5 years prior in Alaska suggests a role for host migration in the spread of these IAVs. Indeed, it is well established that migratory patterns of wild birds contribute substantially to the distribution of the IAV viruses (including high pathogenicity strains like H5) over vast geographic spans.^49–51^ The spread and genetic reassortment of these strains is facilitated by the tendency of multiple migratory shorebirds to congregate in high densities in shared ‘staging’ areas – like Alaska and Iceland – between summer breeding grounds in arctic/subarctic regions and southern wintering areas.^45,51–53^ It is increasingly recognized that targeting surveillance efforts at these hotspots, as well as the migratory routes that stem from them, may be advantageous in tracking viral spread/evolution at the global scale.^53,54^ One intriguing outcome of our study is the notion that passive wastewater surveillance in locations where wildlands abut human settlement might offer this opportunity, increasing overall pandemic preparedness without the need for additional field sampling. However, future work characterizing the specific mechanisms by which wildlife contributes to wastewater, as well as the nature of these contributions, will be necessary to most accurately interpret signals like the ones presented in this study.

## Data Availability

The hybrid-capture metagenomes analyzed here will be made available through the NCBI under BioProject PRJNA1438722. Nucleotide sequences for the assembled influenza segments are available in Table S6. Custom code used to perform the described bioinformatic analyses is available on GitHub: https://github.com/alexanderjaffe/flu-genomics.

## Acknowledgments

We thank Mike Tisza, Alex Crits-Christoph, Marty Freeland, Maya Xu, Katie LaBarbera, and Barbara Mühlemann for helpful discussions and comments on the manuscript. We thank Amanda Bidwell for contributing to the creation of supplementary material. We gratefully acknowledge all data contributors, i.e., the authors and their originating laboratories responsible for obtaining the specimens, and their submitting laboratories for generating the genetic sequence and metadata and sharing via the GISAID Initiative, on which this research is based. Finally, we thank the Palo Alto Regional Water Quality Control Plant, the Santa Clara County Public Health Department, and the California Department of Public Health.

## Funding

This project was supported by a gift from the Sergey Brin Family Foundation to A.B.B.

## Notes

### Competing Interest Statement

The authors have declared no competing interest.

### Summary of Updates

Supplemental tables updated to remove comments from editing process.

