## Supplementary figures/text for "Divergent avian strains drive an off-season influenza A peak in municipal wastewater"

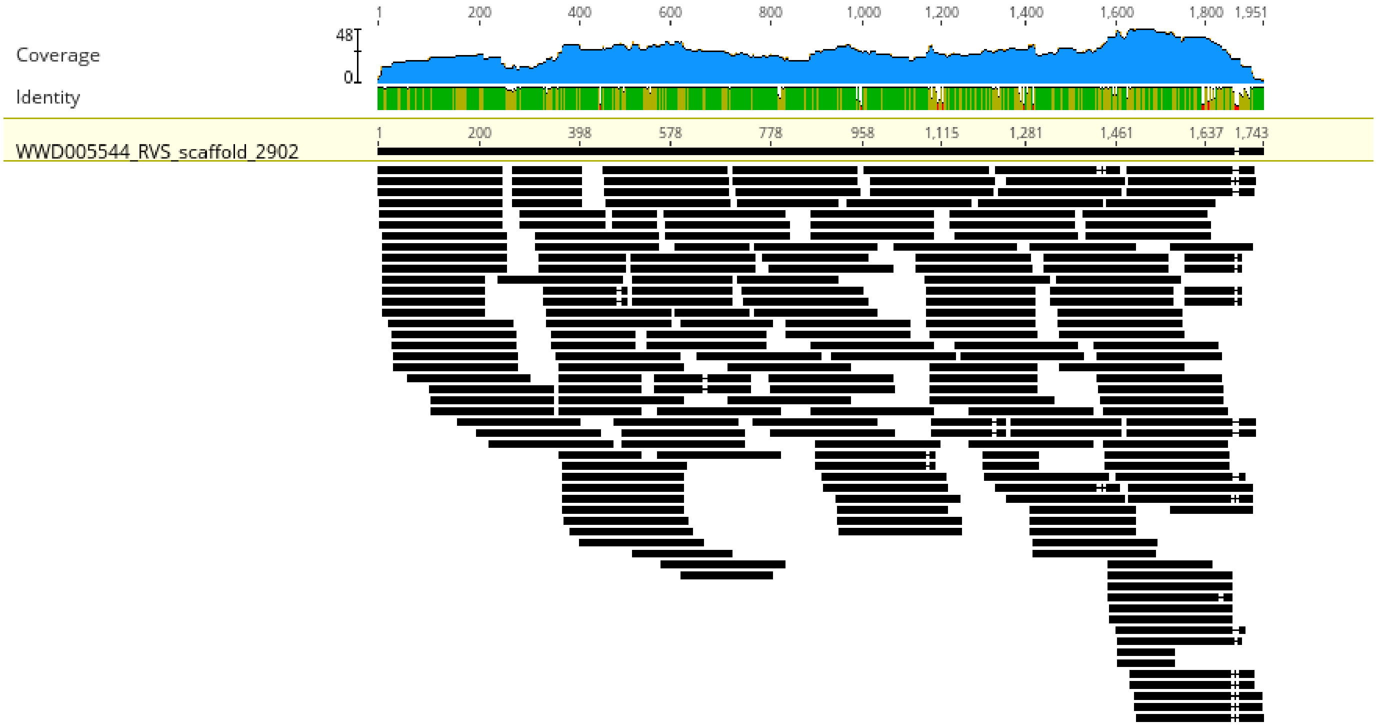


**Figure S1.** Read alignment to an assembled HA segment (WWD005544_RVS_scaffold_2902, length 1,743 bases). Bases are shaded if they agree with the assembled consensus sequence.

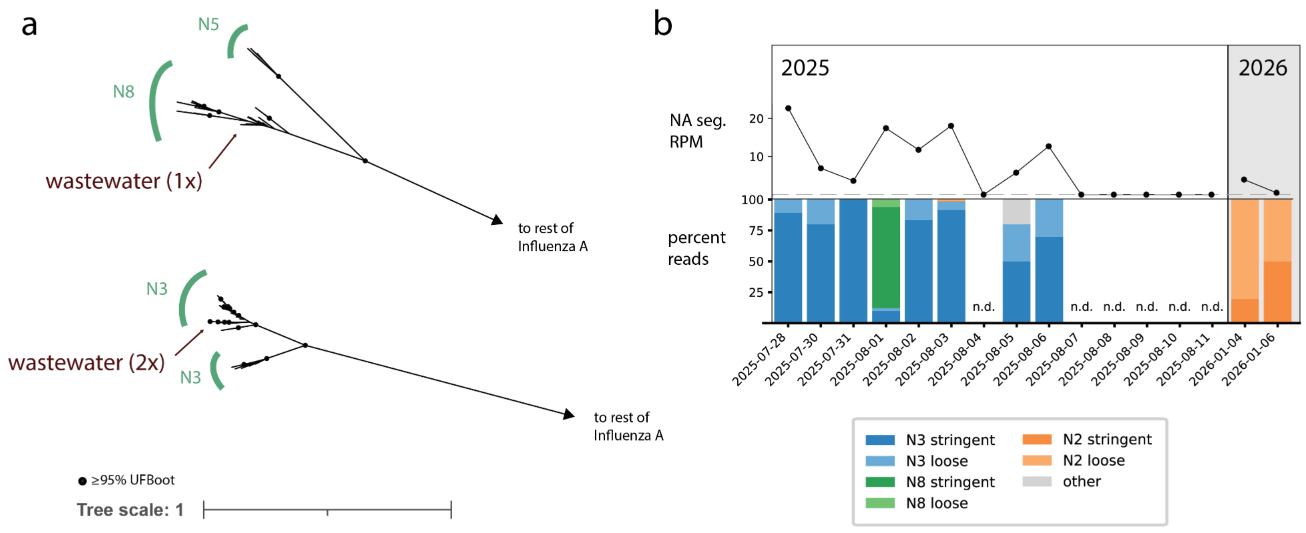


**Figure S2. a)** Subtrees depicting evolutionary relationships of wastewater strains to reference sequences based on the NA protein. Subtrees depict separate regions of the same larger NA protein tree. **b)** Relative abundance dynamics of wastewater strains across the time series, based on reads mapped to a database of NA segments, including newly-assembled sequences. Reads mapping stringently to NA segments, as well as those mapping loosely (Methods) are depicted in different shades. Upper panel indicates the total number of NA reads considered at each time point, expressed in reads per million (RPM). Abbreviations: NA, (neuraminidase); n.d., not detected.

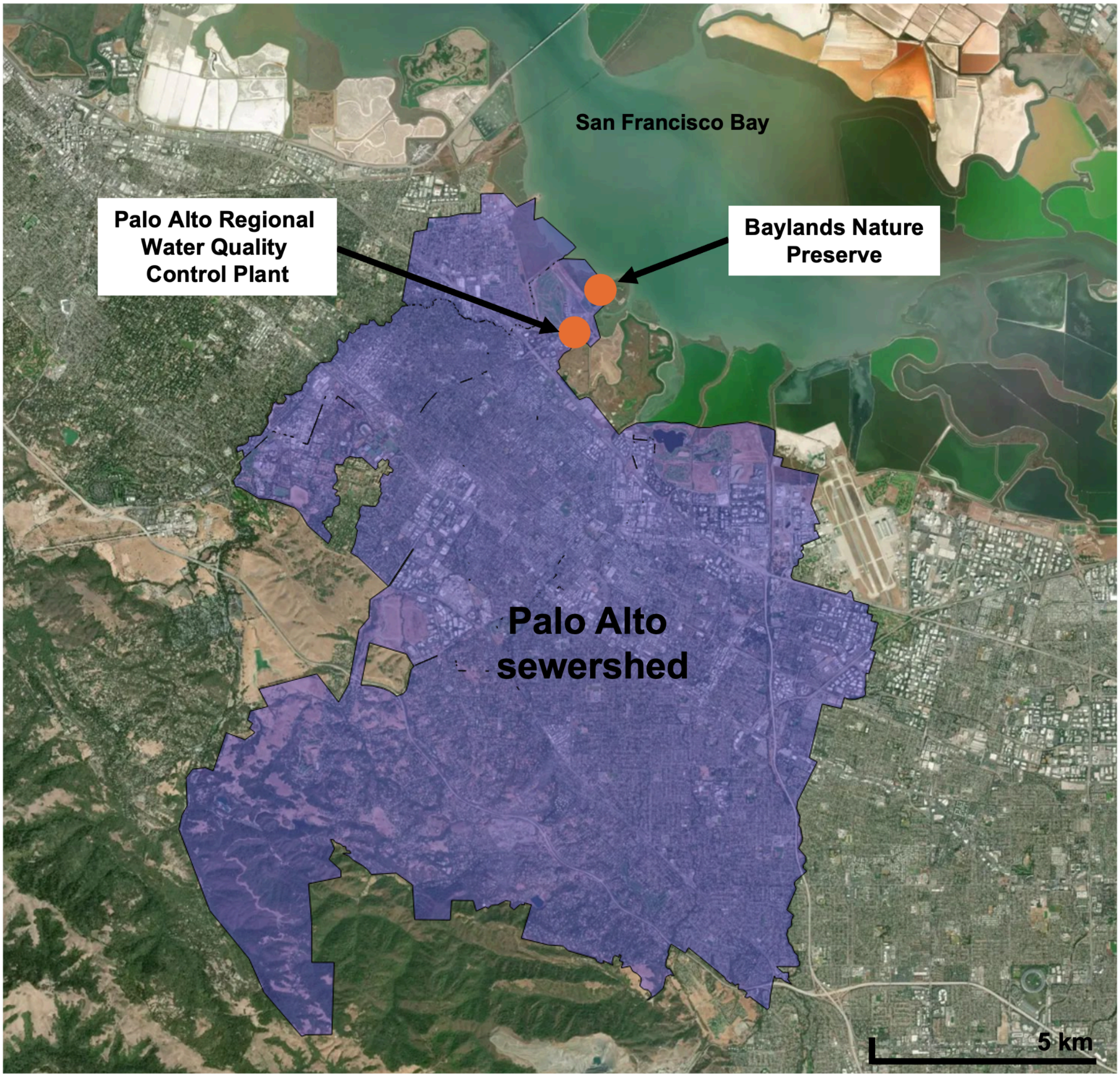


**Figure S3.** Map of the Palo Alto sewershed, including the geographic locations of the Water Quality Control Plant and the Baylands Nature Preserve.

### Supplementary Methods

*In-silico primer/probe specificity testing*

To account for influenza evolution, the specificity and sensitivity of the M, H1, H3, and H5 gene RT-ddPCR assays to circulating IAV subtypes were checked *in silico* monthly during their implementation. At the beginning of each month of the WWSCAN program, the assays for the detection of the Influenza A M gene, H1 gene, H3 gene, and H5 gene are checked *in silico* against the most recently deposited GISAID influenza A sequences. Specifically, for influenza A M gene, sequences from human hosts in the United States for the last 2 months are downloaded, and consensuses are generated at 50%, 75% and 90% consensus. Then, concordance between primers and the resulting consensus sequences is assessed. If there are any mismatches between the two, assays are tested *in vitro* to ensure sensitivity, and updated if major changes occur. For the influenza A Hx genes, sequences from all hosts in the United States for the last 3 months are downloaded, and consensuses are generated at 50%, 75% and 90% consensus. The hemagglutinin gene specific primers are then compared to these consensuses, and mismatches are addressed by updating primers and probes, which then undergo *in vitro* QA/QC testing. Specificity was checked through an exclusionary BLAST that excludes the intended assay target, allowing for the determination of any off target amplification.
